# Mapping Systemic Inflammation and Antibody Responses in Multisystem Inflammatory Syndrome in Children (MIS-C)

**DOI:** 10.1101/2020.07.04.20142752

**Authors:** Conor Gruber, Roosheel Patel, Rebecca Trachman, Lauren Lepow, Fatima Amanat, Florian Krammer, Karen M. Wilson, Kenan Onel, Daniel Geanon, Kevin Tuballes, Manishkumar Patel, Konstantinos Mouskas, Nicole Simons, Vanessa Barcessat, Diane Del Valle, Samantha Udondem, Gurpawan Kang, Sandeep Gangadharan, George Ofori-Amanfo, Adeeb Rahman, Seunghee Kim-Schulze, Alexander Charney, Sacha Gnjatic, Bruce D. Gelb, Miriam Merad, Dusan Bogunovic

**Affiliations:** Precision Immunology Institute, Icahn School of Medicine at Mount Sinai, NY, NY, USA; Mindich Child Health and Development Institute, Icahn School of Medicine at Mount Sinai, NY, NY, USA; Department of Pediatrics, Icahn School of Medicine at Mount Sinai, NY, NY, USA; Department of Microbiology, Icahn School of Medicine at Mount Sinai, NY, NY, USA; Genetics and Genomic Sciences, Icahn School of Medicine at Mount Sinai, NY, NY, USA

**Keywords:** pediatrics, SARS-CoV-2, COVID19, immune, dysfunction, autoimmunity, Kawasaki-like, MIS-C, PIMS

## Abstract

Initially, the global outbreak of COVID-19 caused by severe acute respiratory syndrome coronavirus 2 (SARS-CoV-2) spared children from severe disease. However, after the initial wave of infections, clusters of a novel hyperinflammatory disease have been reported in regions with ongoing SARS-CoV-2 epidemics. While the characteristic clinical features are becoming clear, the pathophysiology remains unknown. Herein, we report on the immune profiles of eight Multisystem Inflammatory Syndrome in Children (MIS-C) cases. We document that all MIS-C patients had evidence of prior SARS-CoV-2 exposure, mounting an antibody response with normal isotype-switching and neutralization capability. We further profiled the secreted immune response by high-dimensional cytokine assays, which identified elevated signatures of inflammation (IL-18 and IL-6), lymphocytic and myeloid chemotaxis and activation (CCL3, CCL4, and CDCP1) and mucosal immune dysregulation (IL-17A, CCL20, CCL28). Mass cytometry immunophenotyping of peripheral blood revealed reductions of mDC1 and non-classical monocytes, as well as both NK- and T-lymphocytes, suggesting extravasation to affected tissues. Markers of activated myeloid function were also evident, including upregulation of ICAM1 and FcγR1 in neutrophil and non-classical monocytes, well-documented markers in autoinflammation and autoimmunity that indicate enhanced antigen presentation and Fc-mediated responses. Finally, to assess the role for autoimmunity secondary to infection, we profiled the auto-antigen reactivity of MIS-C plasma, which revealed both known disease-associated autoantibodies (anti-La) and novel candidates that recognize endothelial, gastrointestinal and immune-cell antigens. All patients were treated with anti-IL6R antibody or IVIG, which led to rapid disease resolution tracking with normalization of inflammatory markers.

**One Sentence Summary:** This study maps the cellular and serological immune dysfunction underlying a novel pediatric inflammatory syndrome associated with SARS-CoV-2.

## Introduction

The rapid spread of severe acute respiratory syndrome coronavirus 2 (SARS-CoV-2) across the globe has led to an outbreak of life-threatening respiratory disease, termed COVID-19 (Zhou et al., 2020; Zhu et al., 2020). While adults have suffered the highest rates of morbidity and mortality of COVID-19, children were thought to be spared (Dong et al., 2020; Ludvigsson, 2020). Recently, however, cases of hyperinflammatory shock in children have been reported in regions with receding SARS-CoV-2 epidemics (Cheung et al., 2020; Jones et al., 2020; Klocperk et al., 2020; Rauf et al.; Riphagen et al., 2020; Toubiana et al., 2020; Verdoni et al., 2020; Whittaker et al., 2020).

Initially, the syndrome was considered an atypical form of Kawasaki disease (KD), an acute systemic vasculitis in young children, given the presence of fever, rash, conjunctivitis, mucocutaneous involvement and cardiac complications (Kawasaki, 1967; Kawasaki et al., 1974). However, it has become evident that shock, gastrointestinal symptoms, and coagulopathy, which are rarely seen in classic KD, are prominent features of this unique syndrome (Cheung et al., 2020; Jones et al., 2020; Klocperk et al., 2020; Rauf et al.; Riphagen et al., 2020; Toubiana et al., 2020; Verdoni et al., 2020; Whittaker et al., 2020). Furthermore, Black and older children appear disproportionately affected, in contrast to the association of young children of Asian descent with KD (Holman et al., 2010; Nakamura et al., 2010). Recognizing these patterns, the World Health Organization (WHO) and other reporting bodies have termed the novel disease multisystem inflammatory syndrome in children (MIS-C) or pediatric inflammatory multisystem syndrome (PIMS) (ECDC, 2020; WHO, 2020).

The concentration of this disease to regions of high local SARS-CoV-2 transmission, but with an onset weeks after the peak COVID-19 caseload, suggests MIS-C is a secondary consequence of SARS-CoV-2 infection. Indeed, over 70% of MIS-C patients test positive for serum antibodies against SARS-CoV-2 and test negative for the presence of viral RNA (Cheung et al., 2020; Jones et al., 2020; Klocperk et al., 2020; Rauf et al.; Riphagen et al., 2020; Toubiana et al., 2020; Verdoni et al., 2020; Whittaker et al., 2020). Aside from this association, the pathophysiology of MIS-C remains largely unexplored. Here, we investigate the immune responses of MIS-C cases, profiling the innate and adaptive underpinnings of the aberrant immune activation.

## Results

### Clinical history

We report eight children from the New York City region that presented to our institution between late-April and early-May 2020 with hyperinflammatory disease fulfilling WHO MIS-C criteria. The median age was 11.5 years, and the gender distribution was equal. Patients that reported ethnicity were of Hispanic (71%) or Black (29%) ancestry. Two patients had a history of asthma and another psychiatric disorders; otherwise, children were previously healthy. All patients initially presented with fever and abdominal symptoms (pain, emesis, or diarrhea). Rash, conjunctivitis, mucocutaneous disease, and hypotension were variably present. None, however, experienced inflammatory manifestations of the extremities, as in KD. On admission, all patients demonstrated signs of coagulopathy as evidenced by elevated fibrin degradation products (D-dimer), PT, PTT and/or thrombocytopenia. Cardiac dysfunction manifested in all patients during hospitalization. Troponin and brain natriuretic protein (BNP) were elevated in all but one patient, with variable electrocardiogram (ECG) changes in three patients. ECG revealed coronary artery dilation or aneurysm in five children. Half of the patients developed respiratory complications, consisting of either reactive airway disease, pleural effusion or pneumonia, although respiratory symptoms were mild. All patients were treated within one day of admission with intravenous immunoglobulin (IVIG), or tocilizumab (TCZ), except P3, for whom IVIG was withheld (Table 1, timeline Supplemental Figure 1A and B). On investigation of SARS-CoV-2 exposure, no patient reported a recent history of upper respiratory infection. When tested during admission, 25% of MIS-C patients were positive by polymerase chain reaction (PCR) for nasopharyngeal SARS-CoV-2 RNA. There was no evidence of other infectious agents. In one of the patients (P4), the mother had a confirmed SARS-CoV-2 infection three weeks prior to admission. Among the patients negative by PCR, one child (P6) had tested positive four weeks previously when he presented with appendicitis. This last case remains the most direct evidence that SARS-CoV-2 infection can precipitate MIS-C weeks later.

**Table 1.** Demographic and clinical features of MIS-C patients. Abbreviations, coronary artery (CA), major depressive disorder (MDD), post-traumatic stress disorder (PTSD), cardiomegaly (CM), mitral regurgitation (MR), left ventricle (LV), thrombocytopenia (TCP), reactive airway disease (RAD).

|  | P1 | P2 | P3 | P4 | P5 | P6 | P7 | P8 | Aggregate |
| --- | --- | --- | --- | --- | --- | --- | --- | --- | --- |
| **Gender** | Male | Female | Male | Female | Male | Male | Female | Female | 50% Male |
| **Comorbidities** | none | none | Asthma | none | Asthma | none | MDD, PTSD | none | 37.50% |
| **WHO MIS-C Criteria** |  |  |  |  |  |  |  |  |  |
| **Fever** | Yes | Yes | Yes | Yes | Yes | Yes | Yes | Yes | 100% |
| **Rash** | No | Yes | No | Yes | Yes | No | No | No | 37.50% |
| **Conjunctivitis** | Yes | Yes | No | No | No | No | No | Yes | 37.50% |
| **Mucocutaneous** | Yes | Yes | No | Yes | No | No | No | No | 37.50% |
| **Extremity** | No | No | No | No | No | No | No | No | 0.00% |
| **Gastrointestinal** | Pain<br>Emesis | Pain<br>Emesis | Pain | Emesis<br>Diarrhea | Diarrhea | Pain | Emesis<br>Diarrhea | Pain<br>Emesis | 100% |
| **Hypotension / shock** | Yes | Yes | No | No | No | Yes | Yes | Yes | 63% |
| **Cardiac Abnormalities** | ↑ troponin<br>↑ BNP<br>CA dilation<br>MR | ↑ troponin<br>↑ BNP | ↑ troponin<br>↑ BNP<br>Prolonged-PR<br>CM | ↑ troponin<br>↑ BNP | CA ectasia | ↑ troponin<br>↑ BNP<br>CA dilation | ↑ troponin<br>↑ BNP<br>CA aneurysm<br>Reduced LV<br>function<br>Diffuse ST<br>elevation | ↑ troponin<br>↑ BNP<br>CA aneurysm<br>Reduced LV<br>function<br>Prolonged QT | 100% |
| **Coagulopathy** | ↑ D-dimer<br>↑ PT<br>↑ PTT | ↑ D-dimer<br>↑ PT<br>↑ PTT | ↑ D-dimer<br>↑ PT<br>↑ PTT | ↑ D-dimer<br>↑ PT<br>↑ PTT | ↑ D-dimer | ↑ D-dimer<br>↑ PT<br>↑ PTT | ↑ D-dimer<br>↑ PT<br>↑ PTT | ↑ D-dimer<br>↑ PT<br>↑ PTT | 87.50% |
| **Inflammatory Markers** | ↑ ESR<br>↑ CRP<br>↑ PCT | ↑ ESR<br>↑ CRP<br>↑ PCT | ↑ ESR<br>↑ CRP | ↑ ESR<br>↑ CRP<br>↑ PCT | ↑ ESR<br>↑ CRP<br>↑ PCT | ↑ ESR<br>↑ CRP<br>↑ PCT | ↑ ESR<br>↑ CRP<br>↑ PCT | ↑ ESR<br>↑ CRP<br>↑ PCT | 100% |
| **Other microbial cause** | No | No | No | No | No | No | No | No | 0% |
| **CoV-2 PCR** | Neg | NA | Neg | Pos | Pos | Neg | Neg | Neg | 25% |
| **Prior Cov-2 Exposure** | NA | NA | NA | Mother PCR+<br>3 weeks prior | NA | PCR+ 4<br>weeks prior | NA | NA | 25% |
| **CoV-2 Serology** | + | + | + | + | + | + | + | + | 100% |
| **Other Clinical Features** | Scrotal pain<br>TCP<br>↑Trans-aminases<br>RAD | TCP<br>↑Trans-aminases<br>RAD | SOB<br>Pleural effusions<br>Thrombocytosis<br>↑Trans-aminases | Thrombocytosis<br>↑Trans-aminases | SOB<br>Headache<br>Hypertension<br>Pneumonia<br>↑Trans-aminases<br>RAD | Appendicitis 4<br>weeks prior<br>↑Trans-aminases | Headache<br>Thrombocytosis<br>↑Trans-aminases | Headache<br>↑Trans-aminases<br>Hypertriglyceridemia |  |
| **Treatment** | IVIG<br>TCZ x3<br>enoxaparin<br>ASA | IVIG x2<br>TCZ x2<br>enoxaparin<br>ASA | TCZ<br>enoxaparin<br>ASA | IVIG x1<br>TCZ x2<br>enoxaparin | IVIG x1<br>TCZ x1<br>enoxaparin | IVIG x1<br>TCZ x2<br>enoxaparin<br>ASA | IVIG x1<br>TCZ x1<br>enoxaparin | IVIG x1<br>TCZ x1<br>enoxaparin<br>ASA<br>hydrocortisone |  |
| **Admission duration (days)** | 8 | 6 | 6 | 5 | 3 | 8 | 6 | 5 | 6 |
| **Outcome** | Favorable | Favorable | Favorable | Favorable | Favorable | Favorable | Favorable | Favorable | Favorable |

Note: All patients are between the ages of 3 - 20

### The anti-SARS-CoV-2 antibody repertoire resembles the convalescent response

Given the suspected association to prior SARS-CoV-2 infection, we performed enzyme-linked immunosorbent assay (ELISA) for presence of serum antibodies against the SARS-CoV-2 spike protein using an FDA-approved protocol (Amanat et al., 2020). All MIS-C patients were seropositive regardless of PCR status (Figure 1A). To better understand the profile of this anti-SARS-CoV-2 response, we explored the isotypes and subclasses of the immunoglobulins specific to SARS-CoV-2 S protein. As a comparator, we included sera from children and adults with acute SARS-CoV-2 infection requiring hospitalization, as well as sera from convalescent adults after mild confirmed infection. Consistent with prior infection, MIS-C plasma showed elevated IgG with low levels of IgM antibody, as observed in the convalscent response. Among the IgG responses, IgG1 predominated, with a detectable but lower IgG3 response—again resembling convalescent sera. Uniquely, however, MIS-C patients showed higher levels of IgA and lower levels of IgM antibodies, relative to convalescent plasma, approximating IgA levels of acute infection (Figure 1A-B). To determine whether this serological response in the absence of clinically apparent respiratory infection was, in fact, effectively antiviral, we assayed neutralization of live SARS-CoV2 infection by patient plasma *in vitro*. All MIS-C patent plasma was capable of neutralization, with potency similar to convalescent reponses. In both ELISA and neutralization assays, PCR^+^ MIS-C patients and PCR^-^ MIS-C patients were indistinguishable (Figure 1C), suggesting that the positive PCR results reflect a receding infection. Indeed, recent studies document that while PCR assays can remain positive beyond three weeks after symptom onset, infectious virus cannot be detected (La Scola et al., 2020; Wölfel et al., 2020; Zheng et al., 2020). To estimate the average time between initial infection and MIS-C onset, we determined the temporal delay between peak COVID-19 and MIS-C admissions at our institution. Our proxy measurement revealed an approximate five week difference (Figure 1D), which is consistent with the documented SARS-CoV2 exposure of P4 and the infection of P6 three and four weeks prior to presentation with MIS-C, respectively.

**Figure 1.**
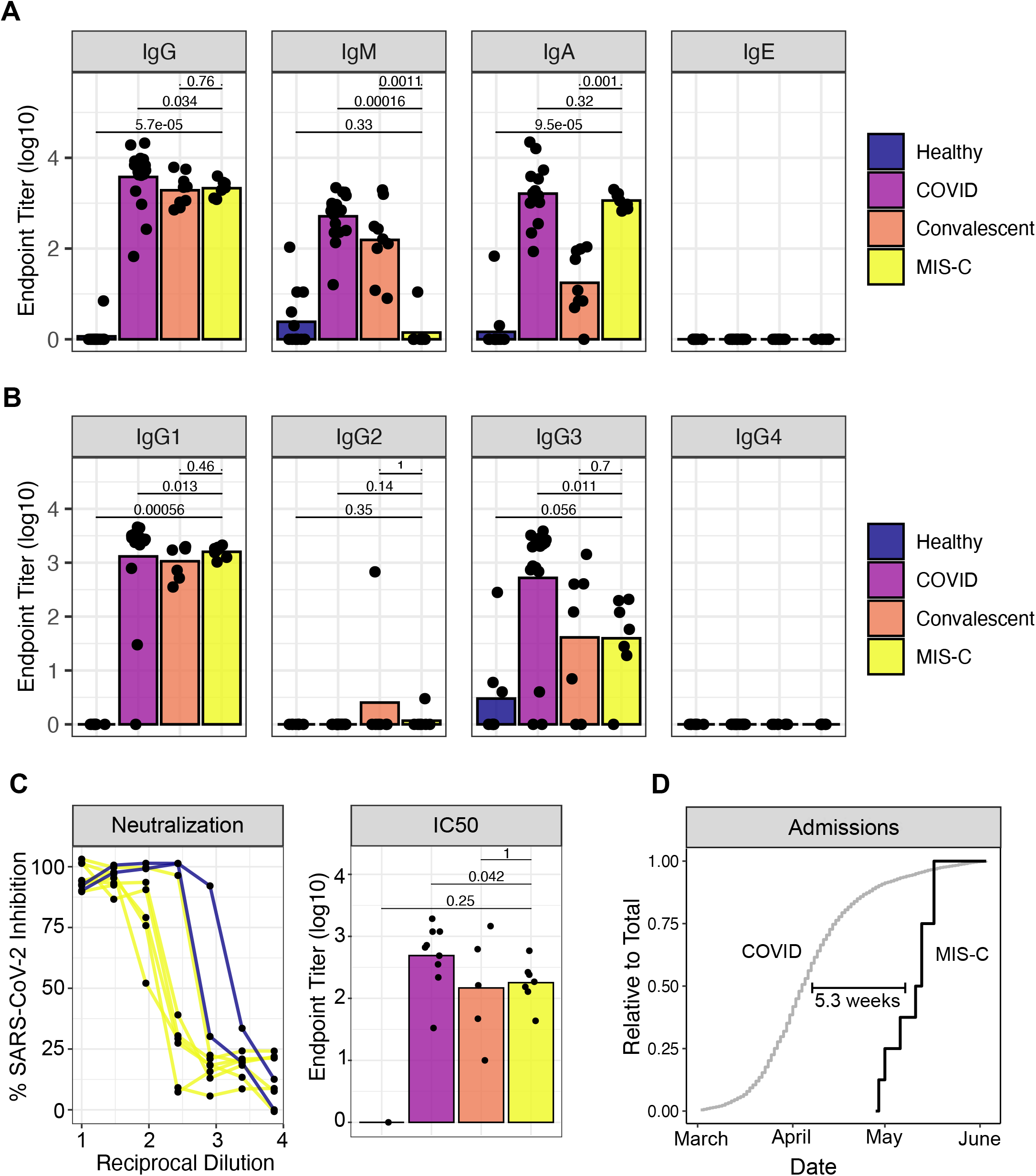
Anti-SARS-CoV-2 humoral response in MIS-C patients. (A) Endpoint titers against SARS-CoV-2 spike protein in plasma from healthy donors, patients hospitalized for acute COVID-19, convalescent individuals, and MIS-C patients. (B) IgG subtypes. (C) Left: Neutralization of SARS-CoV-2 in Vero cells by plasma from MIS-C patients or a representative COVID positive control. Right: IC50 values for neutralization curves across the full dataset. (D) Admissions for COVID-19 and MIS-C, expressed as a relative proportion of total cases, respectively. Time indicates the delay between COVID-19 and MIS-C in the date of 50% total caseload.

### Cytokine profiling indicates myeloid cell chemotaxis and mucosal inflammation

Within a day of admission, all patients received anti-IL6R therapy and all but one received IVIG treatment. We sampled their peripheral blood shortly thereafter, when clinical markers of inflammation, coagulopathy and cardiac dysfunction still remained elevated (Supplemental Figure 1A). We performed high-dimensional cytokine profiling of 92 analytes using the O-link platform to define the secreted immune response in MIS-C patient plasma, and compare it to sera from, age-matched healthy controls, a pool of ten adult healthy donors, or hospitalized pediatric cases of COVID-19 without MIS-C. The latter group included seven pediatric cases (from multiple timepoints) with underlying hemato-oncological disease, and one case of SARS-CoV-2 in an otherwise-healthy child. Overall, the patients with MIS-C presented with striking elevations in multiple cytokine families (Figure 2A). This signature was consistent across patients, as all MIS-C samples grouped together by hierarchical clustering and PCA (Figure 2B). While some of these pro-inflammatory cytokines resembled that of acute SARS-CoV2 infection (IL-18), many of these cytokine elevations exceeded levels of acute infection, such as OPG and IL-10RA (Figure 2C). IL-6 demonstrated the largest fold-change increase, although the exogenous IL-6R blockade from tocilizumab is known to contribute, at least in part, to this effect (Nishimoto et al., 2008).

**Figure 2.**
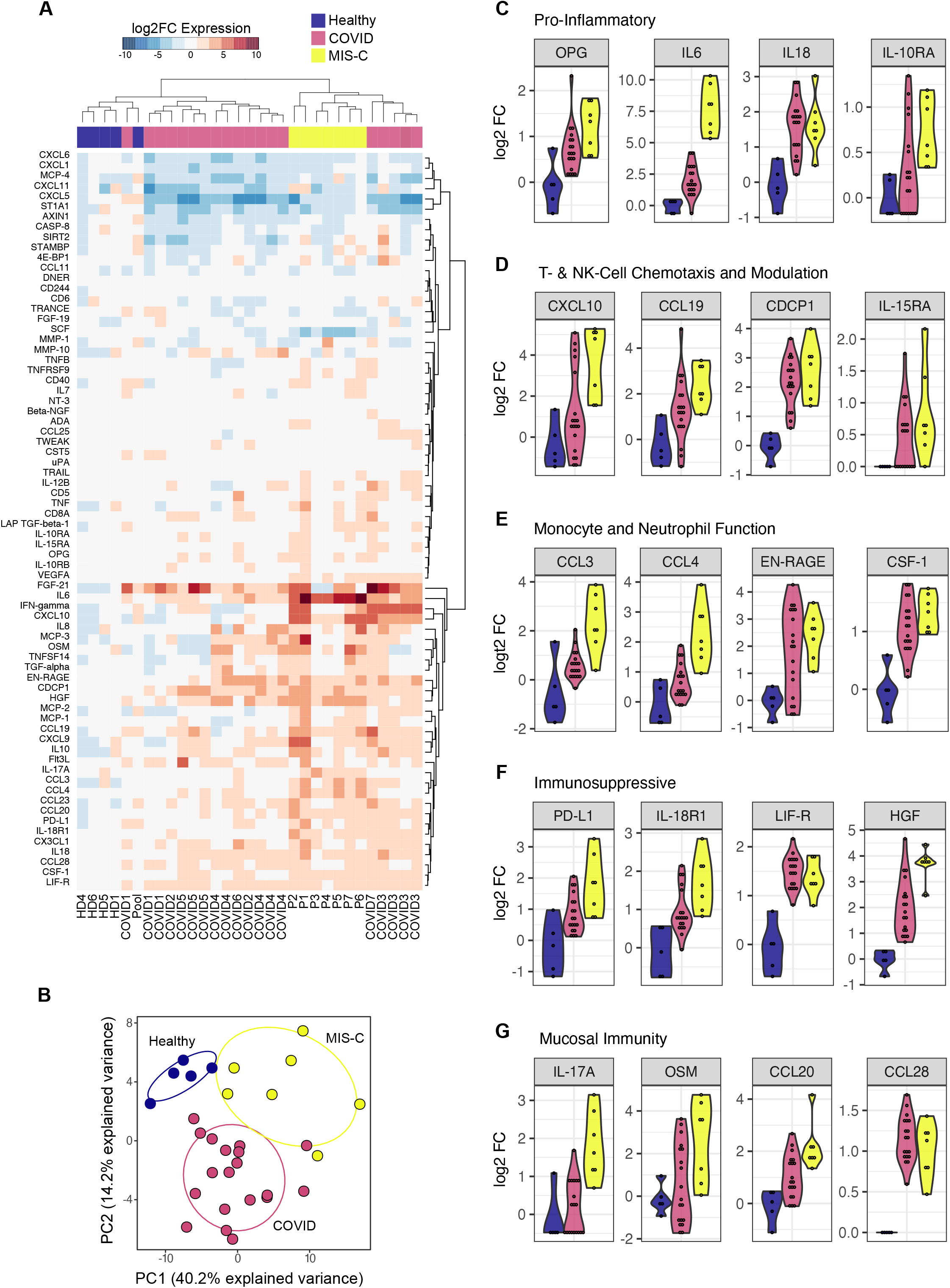
Cytokine profiling of plasma from MIS-C and pediatric COVID-19 patients. (A) Multiplex cytokine analysis by O-link ELISA, expressed as log2 fold-change over the mean healthy controls. (B). Principal component analysis. (C-G) Cytokines reaching statistical significance in MIS-C samples over healthy controls.

Interestingly, the MIS-C circulating immune profile was marked by cytokines and chemokines that recruit NK-cells and T-cells from the circulation and modulate their function such as CCL19, CXCL10 and CDCP1 (Figure 2D) (Vilgelm and Richmond, 2019). Likewise, mediators of neutrophil and monocyte chemotaxis (CCL3 and CCL4), as well as differentiation and activity (EN-RAGE and CSF-1) were elevated in MIS-C (Figure 2E) (Foell et al., 2003; Maurer and Von Stebut, 2004; Stanley and Chitu, 2014; Vilgelm and Richmond, 2019). In turn, a profile of immune exhaustion and suppression was also evident, with stark upregulation of soluble PDL1, likely reflecting a host-driven compensatory response to the inflammation (Figure 2F). In concordance with the gastrointestinal disease of MIS-C, cytokines potentiating mucosal immunity were particularly prominent, both in regards to T-helper cell function (IL-17A) and mucosal chemotaxis (CCL20 and CCL28) (Figure 2F) (Mohan et al., 2020; Williams, 2006). Finally, endothelial dysfunction was suggested by elevation of VEGFA, akin to previous reports in KD (Figure 2A) (Kariyazono et al., 2004; Maeno et al., 1998).

### Mass cytometry uncovers mDC, non-classical monocytes, and lymphocyte activation and egress to the periphery

Next, we performed CyTOF-based immunophenotyping on six MIS-C and five age-matched healthy controls. Overall, while both controls and MIS-C had similar subset distributions in peripheral blood (Figure 3A and Supplemental Figure 2 and 3A), the frequencies of select immune cell types were significantly altered. In the myeloid compartment, mDC1, non-classical monocytes and pDCs were significantly reduced (Figure 3B). Both CD56^bright^ and CD56^dim^ NK cells were also decreased in frequency (Figure 3C), as were the percentages of both αβ and γδ T lymphocytes (Figure 3D). Intrestingly, the relative distribution of naïve, central memory, effector memory or TEMRA subsets was normal within T-cells (Figure 3E and F). Likewise, B-cells were present with a normal frequency range and consisted of a typical distribution of naïve, memory and plasma cells (Supplemental Figure 3B and C). These findings suggests that no active peripheral B- or T-expansion was underway at the time of sampling. Strikingly, the neutrophils and CD16+ monocytes in at least three MIS-C individuals demonstrated robust upregulation of CD54 (ICAM1) expression, indicative of APC activation and transendothelial migration (Figure 3G) (Pietschmann et al., 1998; Sheikh and Jones, 2008). Similarly, MIS-C patient neutrophils and CD16+ non-classical monocytes demonstrated elevated CD64 (FcγR1) expression (Figure 3H), a common finding in autoimmune and autoinflammatory diseases (Li et al., 2009, 2010; Tanaka et al., 2009), including KD (Hokibara et al., 2016). However, these cells lacked signs of active type I IFN signaling, including CD169 and STAT1 phosphorylation upregulation, suggesting other cytokines are driving this activation (Supplemental Figure 3D and E). Instead, augmented levels of phospho-STAT3 were noted, which may originate downstream of IL-6, given its robust elevation in MIS-C plasma (Figure 3I). Combined, these data suggest extravasation of T- and NK-lymphocytes, as well as activation and chemotaxis of neutrophils and nonclassical monocytes, likely contribute to the underlying disease pathogenesis.

**Figure 3.**
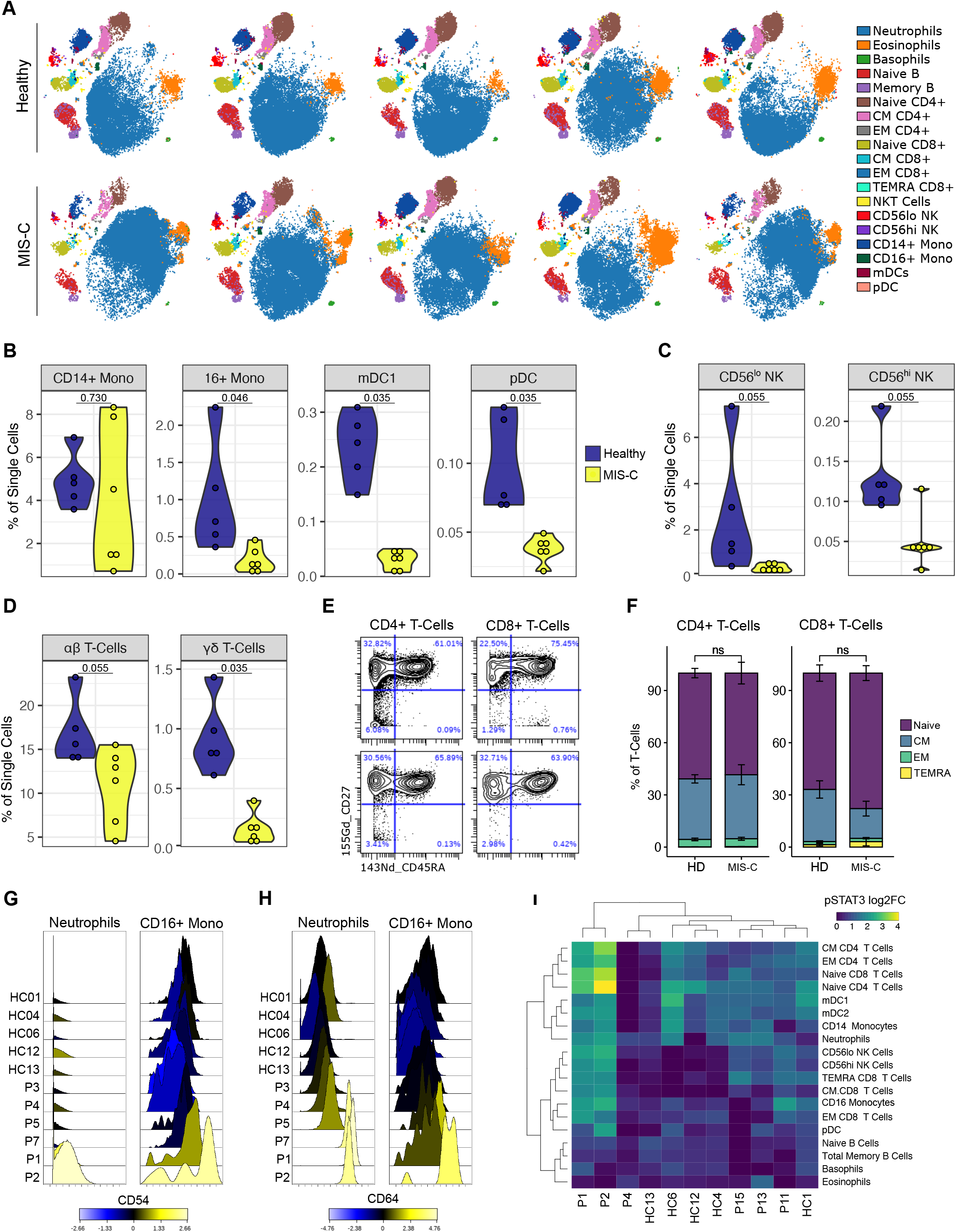
Immunophenotyping of MIS-C patient peripheral blood by mass cytometry. (A) Representative tSNE plots demonstrating the immune cell distribution in whole blood from age-matched healthy controls and MIS-C patients. (B) Monocyte and dendritic cell sub-population frequencies quantified as percent of single cells. (C) NK-cell subsets. (D) T-Cell subsets. (E) Representative scatterplots for naive, central memory (CM), effector memory (EM), and T Effector Memory re-expressing CD45RA (TEMRA) cells in a representative healthy donor (top) and MIS-C patient (bottom). (F) Quantification of T-cell subsets across samples. (G) CD54 and (H) CD64 expression in neutrophils and CD16+ monocytes, color-coded by the log2FC over healthy donor median signal intensity. (I) STAT3 phosphorylation across immune cell subtypes, expressed as log2FC. Abbreviations,

### Identification of auto-antibodies in MIS-C

The resolution of disease with IVIG and the delayed onset after SARS-CoV-2 infection suggest a pathological process involving adaptive immunity. We, therefore, tested the hypothesis that SARS-CoV-2 infection leads to a secondary auto-reactive humoral response. To characterize a potential auto-reactive antibody repertoire, we assessed plasma IgG and IgA reactivity to a human antigen microarray containing over 25,000 unique peptides. To exclude auto-reactive antibodies associated with IVIG treatment (Grüter et al., 2020; Van Der Molen et al., 2015), we first compared plasma from the patient who did not receive IVIG (P3) to plasma from a healthy control and an irrelevant disease control (a multiple myeloma patient). We required a 2-fold change increase in signal intensity between the MIS-C sera and control plasma. We evaluated the global overlap in enriched autoantibody profiles among MIS-C patients (Figure 4A and B). To generate high-confidence hits, we applied a more stringent cutoff (4-fold change) and only assessed auto-reactive antibodies upregulated in the majority of MIS-C patients (Figure 4B and C).

**Figure 4.**
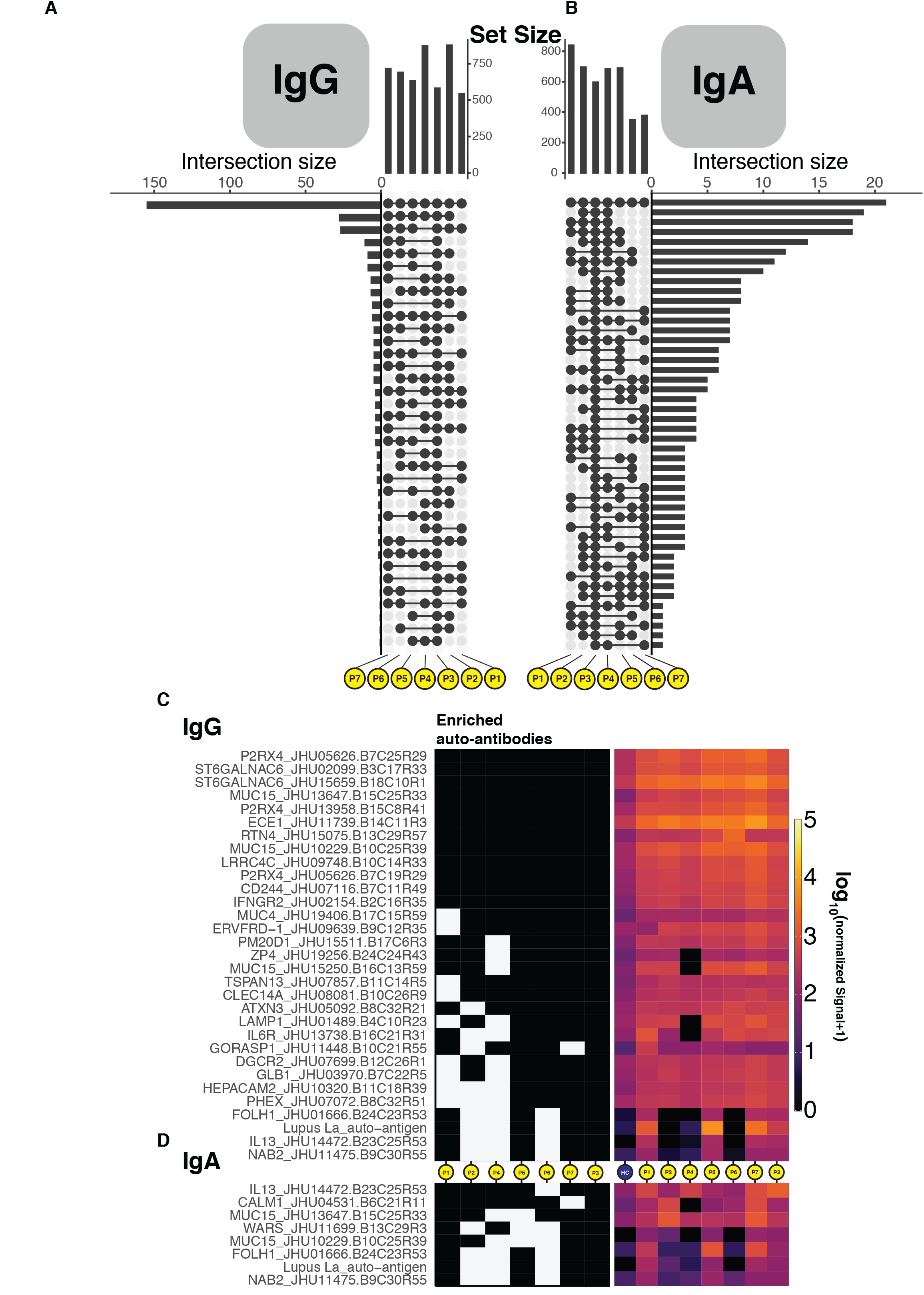
Autoantibody detection unveils an autoreactive repertoire enriched in MIS-C patients. (A-B) Upset plots delineating the number of shared autoantibodies which were at least two-fold enriched when compared to controls for IgG and IgA. (C-D) Tile plots and heatmaps showing the presence or not, and quantile normalized quantity of IgG and IgA autoantigen reactive antibodies that were at least four-fold enriched when compared to controls and found in 4 patients or more, inclusive of patient P3.

This analysis yielded 25 candidates for IgG autoantigens and 7 for IgA autoantigens (Figure 4C and D). Among this group, anti-La, a characteristic autoantigen of systemic lupus erythematosus and Sjogren’s disease, was enriched in both IgG and IgA libraries as compared to controls (Franceschini and Cavazzana, 2005). Likewise, IgG reactivity to aminoacyl-tRNA synthetase (WARS) were identified, similar to that observed in polymyositis and dermatomyositis (Mirrakhimov, 2015). These two candidates suggest that MIS-C may share some pathophysiology with classic autoimmune diseases. Interestingly, most auto-reactive peptides noted to be enriched in our MIS-C patients carry no documented association with autoimmune disease. The tissue expression patterns of these antigens reveal enrichment in organ systems central to the pathology of MIS-C. Among these were peptides expressed in endothelial and cardiac tissue (P2RX4, ECE2 and CLEC14A), as well as antigens of the gastrointestinal tract (MUC4, MUC15 and FOLH1). Curiously, immune cell mediators were particularly abundant, including CD244, IL13, IFNGR2, IL6R, and LAMP1. Whether antibody engagement with these proteins modulates the activity of immune cells or aids in immune complex formation needs to be determined. Plausibly, antibody-mediated inhibition of CD244, an inhibitory receptor on NK and T-Cells, could allow for breach of immune checkpoints. Future studies are required to confirm this association in MIS-C cohorts and assess the functional impact.

### Anti-IL-6R therapy and IVIG resolve MIS-C

Beginning on the day of admission, we monitored markers of inflammation (C-reactive protein, erythrocyte sedimentation rate, IL-6, IL-8, IL1-beta, TNF-alpha, ferritin), coagulopathy (D-dimer, prothrombin time, partial thromboplastin time, platelet count, fibrinogen) and cardiac injury (troponin and BNP). Most patients were treated within the first day of admission. All received anti-IL-6R antibody, and all but one received IVIG. Uniformly, these markers normalized rapidly (Supplemental Figure 1 and 2) with a median hospital stay of six days, and favorable outcomes. We continued to monitor these disease parameters on follow-up, noting that they continued to normalize without evident secondary consequences.

## Discussion

Here, we evaluated the peripheral blood immune profiles of eight MIS-C cases. Despite the absence of clinically apparent upper respiratory infection, all children harbored antibodies against SARS-CoV-2. This antibody response demonstrated typical IgG class switching, loss of circulating IgM, and effective virus neutralization, resembling, but not identical to, serologies from convalescent COVID-19 adults. Their peripheral blood secretome exhibited drastic elevations of inflammatory mediators, indicative of lymphocyte and myeloid cell activation and chemotaxis towards the periphery. Among these cytokines, a clear mucosal immune signature was evident, in accordance with the prominent gastrointestinal clinical manifestations. Cellular analyses supported egress of nonclassical monocytes and mDC1s, as well as T and NK cells, from the periphery. Importantly, we identified IgG and IgA auto-antibody repertoires against endothelial, mucosal and immune antigens, together with strong neutrophil and monocyte upregulation of CD54 and CD64. The latter marker, also known as the high-affinity FcγR1, can engage autoantibodies and immune complexes to trigger potent inflammation and tissue injury (van der Poel et al., 2011; Tanaka et al., 2009). These results suggest that autoreactivity secondary to SARS-CoV-2 infection and the inflammatory innate immune response may be critical to the pathogenesis of MIS-C.

In 1967, Dr. Tomisaku Kawasaki described 50 pediatric patients with a previously unrecognized febrile illness that clustered both in time and geography (Kawasaki, 1967). Since this initial description, numerous studies have detailed the clinical features and biological manifestations of KD (Dietz et al., 2017). However, the underlying pathophysiology remains incompletely understood. Most mechanistic explanations arise from the association with viral infections. Namely, there is a significant increase in the incidence of PCR-positive tests for enteroviral, adenoviral, human rhinoviral and coronaviral infections in children presenting with KD relative to age-matched, healthy controls (Chang et al., 2014; Jordan-Villegas et al., 2010; Turnier et al., 2015). Likewise, according to serologic and epidemiologic evidence, we observed that all MIS-C patients were previously exposed to SARS-CoV-2 approximately four to five weeks prior to presentation. While MIS-C has been classified as a distinct syndrome by its clinical presentation, the overlapping features are striking, suggesting that MIS-C may lie along a spectrum of KD-like pathology. These differences may arise from the introduction of a novel virus to a population with completely naïve immunity, as in SARS-CoV-2. This distinction may underlie the later age at presentation for MIS-C relative to KD, as other viruses associated with KD are common infections of early childhood. Should a different experiment of nature have occurred whereby other KD-associated viruses suddenly appeared in a naïve population, it is plausible that distinct clinical and laboratory features would also have manifested, linked to those viruses.

The extent to which genetics impacts the development of MIS-C is currently unclear. Certainly, Black or Hispanic ethnicity appears to be a major risk factor, as observed in this study and others (Cheung et al., 2020; Jones et al., 2020; Klocperk et al., 2020; Rauf et al.; Riphagen et al., 2020; Toubiana et al., 2020; Verdoni et al., 2020; Whittaker et al., 2020). This enrichment diverges from KD, in which the incidence is significantly higher in children of Asian ancestry. Several genetic variants of moderate effect size, such as ITPKC, CD40, FCGR2A and BLK have been associated with KD (Onouchi, 2018). Interestingly, the risk among Asian children living in the United States is reduced (Uehara and Belay, 2012), suggesting a role for environmental factors. Similarly, it is quite possible that environmental factors impact the ethnicity-based risk in MIS-C, as Black and Hispanic populations are more likely to be exposed to SARS-CoV2 (DiMaggio et al., 2020; Vahidy et al., 2020). This factor is especially relevant at our hospital, and other metropolitan centers, which serve patients from diverse backgrounds. Only detailed genetic analyses in large cohorts will determine the relative contribution of genetic factors, which in KD also remains mostly unexplained.

Recurrence of KD is rare, and hopefully this will be the case for MIS-C as well. The presence of autoantibodies, as documented here, is concerning however. We postulate that these autoantibodies trigger immune complex formation and/or unleash an immune cell-driven attack against host tissues. These may arise by direct cross-reactivity between SARS-CoV-2 and self-antigens, which, if true, will pose a risk for future vaccination strategies. Although the inflammation appears transient, these autoantibodies also raise concern for recurrence or predisposition to other disorders with autoimmune features. All these postulates need careful, methodical and well-controlled experimental dissection. Until then, MIS-C remains scientifically puzzling, but therapeutically manageable.

## Data Availability

Data will be made available upon request to the authors.

## Acknowledgments

This research was supported by National Institute of Allergy and Infectious Diseases Grants R01AI127372, R21 AI134366 and R21AI129827, and funding from the March of Dimes, awarded to DB. CG was supported by T32 training grant 5T32HD075735-07. S.G., D.M.D.V.. S.K.-S., A.R. and M.M. were supported by NCI U24 grant CA224319. S.G. is additionally supported by grants U01 DK124165, P01 CA190174, PCF Challenge Award and DOD W81XWH-18-1-0528. M.M. was supported by the fast-grant fund. The Human Immune Monitoring Center and the Institute for Healthcare Delivery Science received support from Cancer Center P30 grant CA196521.

## STAR Methods

### Sample collection

Written informed consent for all individuals in this study was provided in compliance with an institutional review board protocol. Acutely-ill and convalescent patients were recruited at the Mount Sinai Health System. Healthy volunteers were age-matched to the extent possible, including a 3 year-old female, 6 year-old female, 7 year old male, 12 year-old male, and 19 year-old female. For cytokine assays, an additional healthy volunteer pool was generated from adult sera. From each patient, blood was drawn into a Cell Preparation Tube with sodium heparin (BD Vacutainer) and serum separating tubes (SST) processed immediately. Whole blood was fixed using Proteomic Stabilizer PROT1 (SmartTube) and frozen at −80C. Peripheral blood mononuclear cells and plasma/sera was isolated by centrifugation and subsequently stored at - 80°C until use.

### Serology and neutralization

The development and protocol for the SARS-CoV-2 spike antigen is described elsewhere in detail (Amanat 2020). Briefly, sera from each time point were tested in each patient using serial 4 × dilutions from 1/100 to 1/6,400 for reactivity to full-length SARS-CoV-2 recombinant protein (0.5 µg/mL). Titers were extrapolated based on a cutoff established from a pool of healthy donor sera, and assays were validated using positive control sera for each antigen present on each plate. Results were considered significant if titers were ≥ 100. To assess the distribution of different immunoglobulin isotypes, assays were performed separately with anti-human IgA-AP antibody, anti-human IgM-AP antibody, anti-human IgG1 Fc-AP, anti-human IgG2 Fc-AP, anti-human IgG3 hinge-AP and anti-human IgG4 Fc-AP. Endpoint titers were calculated by the last dilution before reactivity dropped below an optical density threshold defined by the OD of a healthy donor pool.

### Microneutralization assay

Heat-inactivated plasma samples were serially diluted in complete media (10% 10× minimal essential medium (Gibco), 2 mM L-glutamine, 0.1% sodium bicarbonate (wt/vol; Gibco), 10 mM 4-(2-hydroxyethyl)-1-piperazineethanesulfonic acid (HEPES; Gibco), 100 U ml–1 penicillin, 100 ug/ml–1 streptomycin (Gibco) and 0.2% bovine serum albumin (MP Biomedicals). Diluted plasma was then incubated in a 1:1 volumetric ratio with SARS-CoV-2 virus (USA-WA1/2020; GenBank: MT020880) at a concentration of 100 TCID50 (50% tissue culture infectious dose) in 1× MEM for 1 hour at room temperature. This virus–serum mixture was then added to Vero E6 cells (ATCC) plated in a 96-well cell culture plate and incubated at 37 °C for 1 hour. The supernatant was then removed, and the diluted plasma samples were re-added for 48 hours at 37 °C. The infected cells were then fixed with 10% paraformaldehyde (Polysciences) for 24 hours at 4 °C. Following fixation, the cells were washed, permeabilized with 0.1% Triton X-100, blocked in a 3% milk solution (American Bio) and stained with a monoclonal antibody to anti-SARS nucleoprotein (Amanat 2020) and subsequently a goat anti-mouse IgG–HRP (Rockland Immunochemicals). A reaction with SIGMAFAST OPD (Sigma–Aldrich) was carried out and the OD at 490nm was measured. A threshold value of the mean optical density value of blank wells plus three standard deviations was established and used to determine the microneutralization titer. Microneutralization assays were performed in a facility with a biosafety level of 3.

### Multiplex cytokine analysis

For analysis of circulating cytokines, we used the O-link proteomics inflammation panel, which consists of 92 inflammation-related proteins quantified by an antibody-mediated proximity extension-based assay. Analytes with normalized protein expression values below the limit-of-detection in >75% of samples were excluded from further analysis. For the remainder of analytes, any sample under the limit of detection was assigned a value of the limit-of-detection divided by the square root of 2. The log2 fold-change over the median healthy control protein expression was then calculated, and the Benjamini-Hochberg procedure was used to adjust P values for multiple testing.

### Mass cytometry

Frozen stabilized blood samples were thawed according to the manufacturer’s recommended protocol, then washed with barcode permeabilization buffer (Fluidigm). Samples were uniquely barcoded with Cell-ID 20-Plex Pd Barcoding Kit (Fluidigm), washed and pooled together. An Fc-block and a heparin-block were then performed to prevent non-specific binding. Cells were then incubated with an antibody cocktail for surface markers to identify major immune populations. All antibodies were purchased with pre-conjugated or conjugated in-house with X8 MaxPar conjugation kits (Fluidigm). After surface staining, the samples were methanol-permeabilized, washed, heparin-blocked and stained with a cocktail of antibodies against intracellular targets, including markers of phosphorylation and signaling. After washing, cells were then incubated in freshly diluted 2.4% formaldehyde containing 125nM Ir Intercalator (Fluidigm), 0.02% saponin and 30 nM OsO4 (ACROS Organics) for 30 minutes at room temperature. Samples were then washed and acquired immediately.

For acquisition, samples were washed with PBS+0.2% BSA, PBS, and then CAS buffer (Fluidigm). The final solution in CAS buffer consisted of 1 million cells per mL and a 1/20 dilution of EQ beads (Fluidigm). Following routine instrument optimization, samples were acquired at a rate of <300 events per second on a Helios mass cytometer (Fluidigm) with a modified wide-bore injector (Fluidigm). FCS files were then normalized and concatenated with Fluidigm acquisition software, and deconvoluted with a Matlab-based debarcoding application (“*Palladium-based mass tag cell barcoding with a doublet-filtering scheme and single-cell deconvolution algorithm*”).

FCS files were analyzed using Cytobank. Cell events were identified as Ir191/193-positive and Ce140-negative events. Doublets were excluded on the basis of Mahalanobis distance and barcode separation and with the Gaussian parameters acquired with Helios CyTOF software. Downstream data analysis was performed on Cytobank, by both tSNE analysis and biaxial gating of immune populations. Statistical significance for cell population frequencies between healthy controls and patients were determined by the Benjamini-Hochberg procedure to correct for multiple testing.

### Auto-antibody specificity analysis

Seromic profiling of autoantibodies was conducted as previously described (Gnjatic et al., 2010). These assays used the CDI HuProt array. Seven MIS-C plasma samples and 3 additional controls samples were run in parallel. CDI digital image output was manually aligned and flagged for artifacts. To minimize non-specific, background signal, median fluorescent values were calculated and subtracted from digital images. Final signal intensities were determined by replicate averaging and a quantile normalization was applied. Due to disparity of computational packages dedicated protein microarray analysis, we treated quantile normalized data in an approach akin to RNA-chip microarray analysis. Quantile normalized matrices were read into R statistical environment (4.0.1) and analyzed using the DESeq2 and limma R packages. Pair-wise contrasts between each MIS-C patient versus the controls and an irrelevant disease control were conducted and differentially enriched antigen lists were generated. To control for noise and filter low intensity signals, antigens required to have a normalized signal intensity of 86 (the 75^th^ percentile of total signal) or greater in at least one patient to be tested. Lists were further filtered to only include antigens that exhibited a 4-fold enrichment compared to control samples for heatmap visualization. Overlap of enriched samples were visualized by the UpSetR package and tile plot and heat map visualizations were drawn by the ggplot2 software. All code and scripts will be made available upon request.

## Supplementary Information

## Supplementary Figure Legends

**Figure S1 (Related to Table 1).**
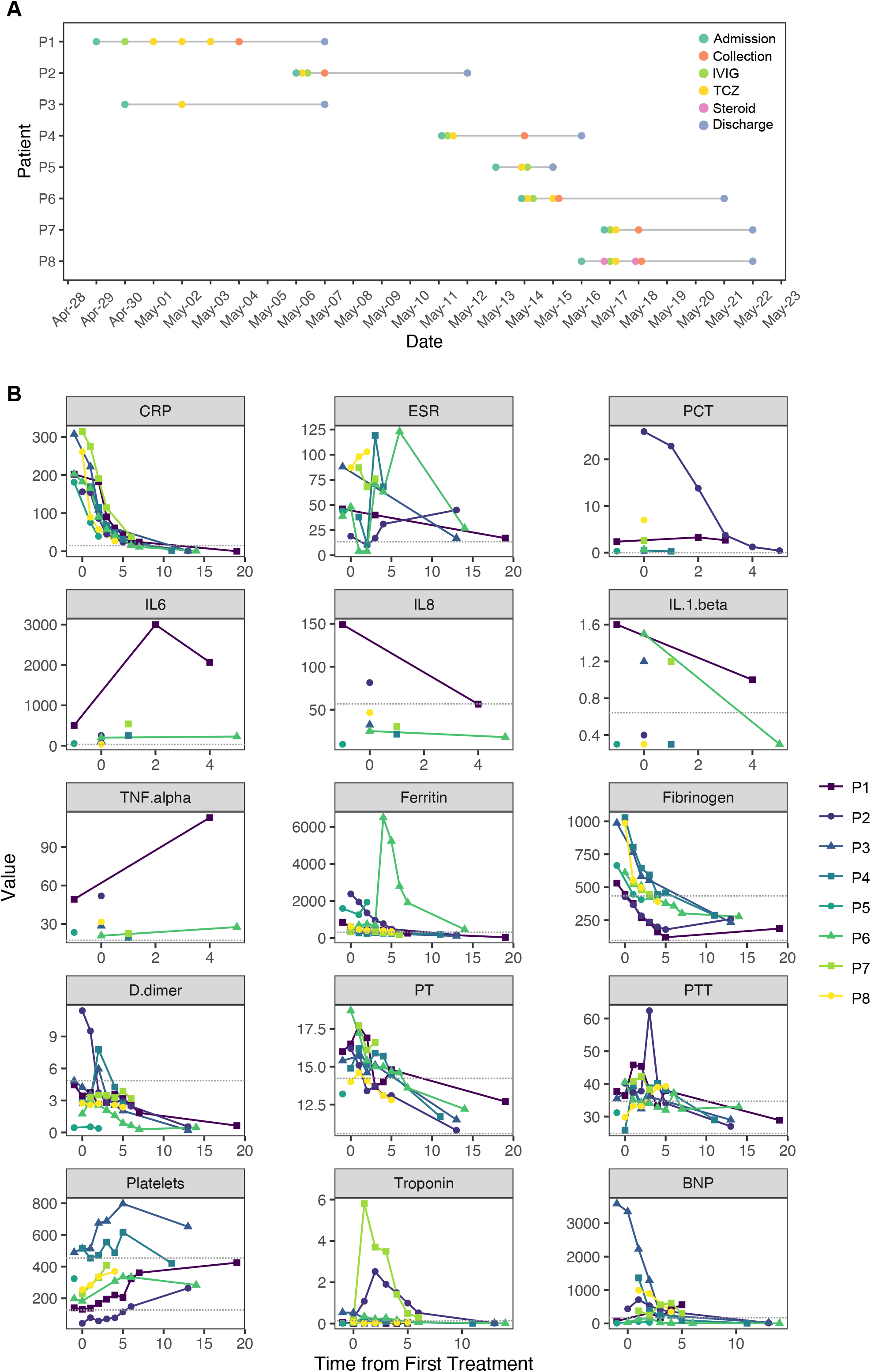
MIS-C disease course and sample collection. (A) Timeline of admission, treatment, sample collection and discharge. (B) Laboratory markers of inflammation, coagulopathy and cardiac dysfunction in response to IVIG and/or TCZ treatment. Dotted lines represent normal limits.

**Figure S2 (Related to Table 1).**
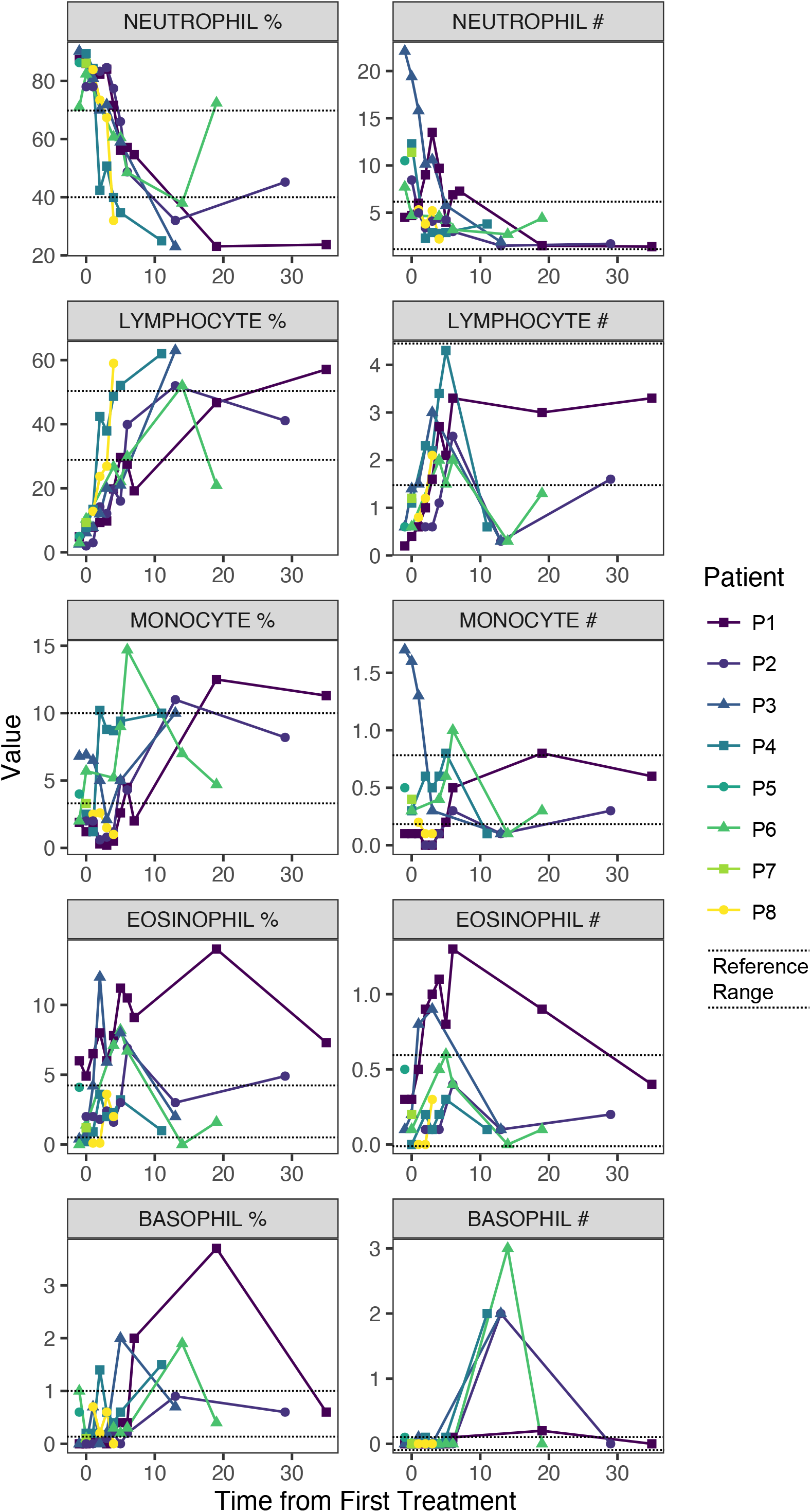
White blood cell count and differential in response to TCZ and IVIG treatment. (Left) Percentage values of immune cell types during treatment course to TCZ and IVIG across all MIS-C patients. (Right) Absolute counts of immune cell types during treatment course to TCZ and IVIG across all MIS-C patients. Dotted lines represent normal limits.

**Figure S3 (Related to Figure 3).**
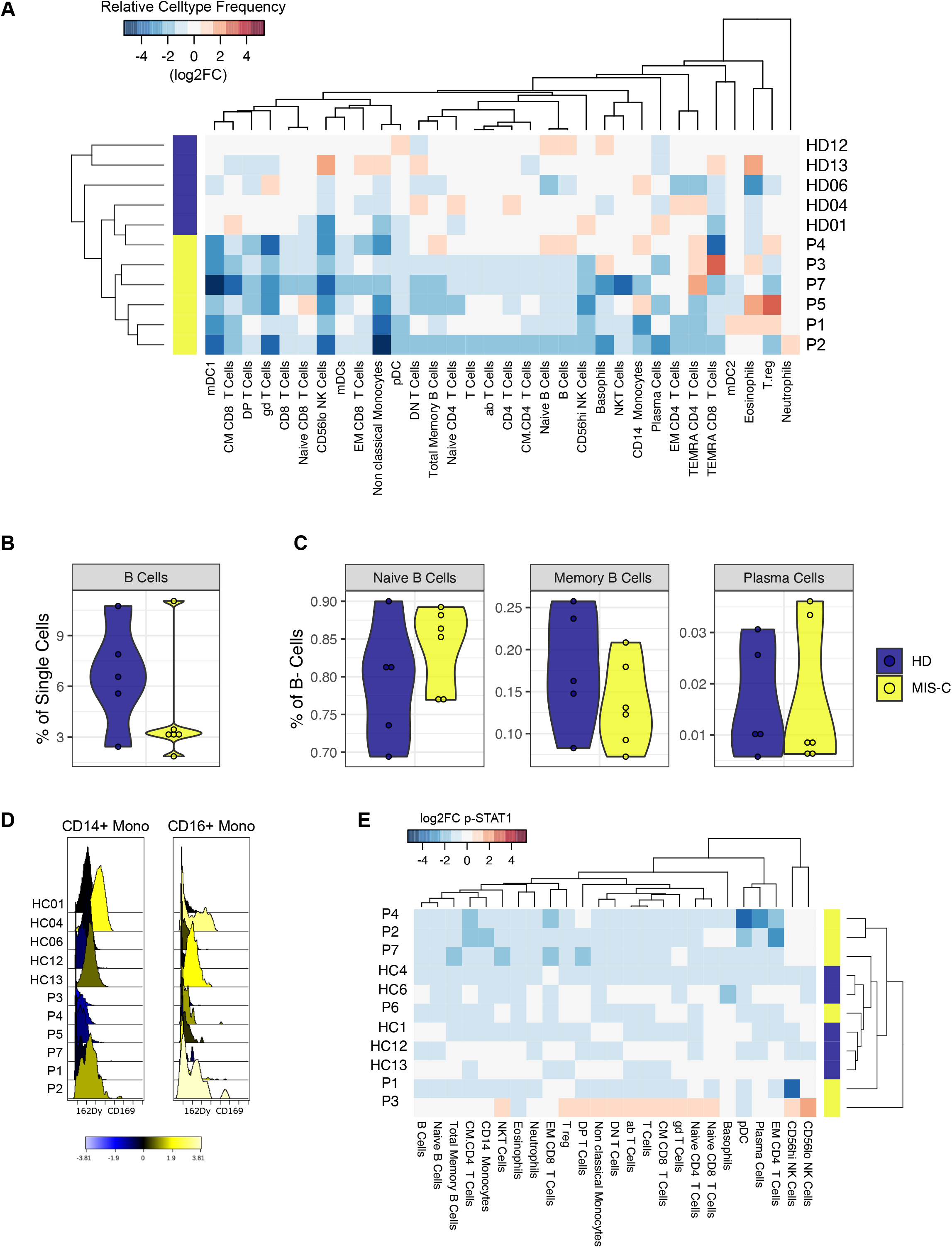
Mass cytometry of peripheral blood immune cells. (A) Immune cell frequencies expressed as the log2 fold change over the mean of healthy controls. (B). B-cell frequency as percent of single cells. (C). Expression of CD169, an interferon-stimulated gene, in monocytes. (E) Relative STAT1 phosphorylation.

## References

Amanat, F., Stadlbauer, D., Strohmeier, S., Nguyen, T.H.O., Chromikova, V., McMahon, M., Jiang, K., Arunkumar, G.A., Jurczyszak, D., Polanco, J., et al. (2020). A serological assay to detect SARS-CoV-2 seroconversion in humans. Nat. Med.

Chang, L.-Y., Lu, C.-Y., Shao, P.-L., Lee, P.-I., Lin, M.-T., Fan, T.-Y., Cheng, A.-L., Lee, W.-L., Hu, J.-J., Yeh, S.-J., et al. (2014). Viral infections associated with Kawasaki disease. J. Formos. Med. Assoc. 113, 148–154.

Cheung, E.W., Zachariah, P., Gorelik, M., Boneparth, A., Kernie, S.G., Orange, J.S., and Milner, J.D. (2020). Multisystem Inflammatory Syndrome Related to COVID-19 in Previously Healthy Children and Adolescents in New York City. JAMA 8–10.

Dietz, S.M., van Stijn, D., Burgner, D., Levin, M., Kuipers, I.M., Hutten, B.A., and Kuijpers, T.W. (2017). Dissecting Kawasaki disease: a state-of-the-art review. Eur. J. Pediatr. 176, 995–1009.

DiMaggio, C., Klein, M., Berry, C., and Frangos, S. (2020). Blacks/African Americans are 5 Times More Likely to Develop COVID-19: Spatial Modeling of New York City ZIP Code-level Testing Results. MedRxiv 2020.05.14.20101691.

Dong, Y., Mo, X., Hu, Y., Qi, X., Jiang, F., Jiang, Z., and Tong, S. (2020). Epidemiology of COVID-19 Among Children in China. Pediatrics 145, e20200702.

ECDC (2020). Paediatric inflammatory multisystem syndrome and SARS-CoV-2 infection in children. 1–18.

Foell, D., Kucharzik, T., Kraft, M., Vogl, T., Sorg, C., Domschke, W., and Roth, J. (2003). Neutrophil derived human S100A12 (EN-RAGE) is strongly expressed during chronic active inflammatory bowel disease. Gut 52, 847–853.

Franceschini, F., and Cavazzana, I. (2005). Anti-Ro/SSA and La/SSB antibodies. Autoimmunity 38, 55–63.

Gnjatic, S., Ritter, E., Büchler, M.W., Giese, N.A., Brors, B., Frei, C., Murray, A., Halama, N., Zörnig, I., Chen, Y.T., et al. (2010). Seromic profiling of ovarian and pancreatic cancer. Proc. Natl. Acad. Sci. U. S. A. 107, 5088–5093.

Grüter, T., Ott, A., Meyer, W., Jarius, S., Kinner, M., Motte, J., Pitarokoili, K., Gold, R., Komorowski, L., and Ayzenberg, I. (2020). Effects of IVIg treatment on autoantibody testing in neurological patients: marked reduction in sensitivity but reliable specificity. J. Neurol. 267, 715–720.

Hokibara, S., Kobayashi, N., Kobayashi, K., Shigemura, T., Nagumo, H., Takizawa, M., Yamazaki, T., and Agematsu, K. (2016). Markedly elevated CD64 expression on neutrophils and monocytes as a biomarker for diagnosis and therapy assessment in Kawasaki disease. Inflamm. Res. 65, 579–585.

Holman, R.C., Belay, E.D., Christensen, K.Y., Folkema, A.M., Steiner, C.A., and Schonberger, L.B. (2010). Hospitalizations for Kawasaki Syndrome Among Children in the United States, 1997–2007. Pediatr. Infect. Dis. J. 29, 1.

Jones, V.G., Mills, M., Suarez, D., Hogan, C.A., Yeh, D., Bradley Segal, J., Nguyen, E.L., Barsh, G.R., Maskatia, S., and Mathew, R. (2020). COVID-19 and Kawasaki Disease: Novel Virus and Novel Case. Hosp. Pediatr.

Jordan-Villegas, A., Chang, M.L., Ramilo, O., and Mejías, A. (2010). Concomitant respiratory viral infections in children with kawasaki disease. Pediatr. Infect. Dis. J. 29, 770–772.

Kariyazono, H., Ohno, T., Khajoee, V., Ihara, K., Kusuhara, K., Kinukawa, N., Mizuno, Y., and Hara, T. (2004). Association of vascular endothelial growth factor (VEGF) and VEGF receptor gene polymorphisms with coronary artery lesions of Kawasaki disease. Pediatr. Res. 56, 953–959.

Kawasaki, T. (1967). [Acute febrile mucocutaneous syndrome with lymphoid involvement with specific desquamation of the fingers and toes in children]. Arerugi 16, 178–222.

Kawasaki, T., Kosaki, F., Okawa, S., Shigematsu, I., and Yanagawa, H. (1974). A new infantile acute febrile mucocutaneous lymph node syndrome (MLNS) prevailing in Japan. Pediatrics 54, 271–276.

Klocperk, A., Parackova, Z., Dissou, J., Malcova, H., Pavlicek, P., Vymazal, T., Dolezalova, P., and Sediva, A. (2020). Systemic inflammatory response and fast recovery in a pediatric patient with COVID-19. Res. Prepr. 1–6.

Li, Y., Lee, P.Y., Sobel, E.S., Narain, S., Satoh, M., Segal, M.S., Reeves, W.H., and Richards, H.B. (2009). Increased expression of FcγRI/CD64 on circulating monocytes parallels ongoing inflammation and nephritis in lupus. Arthritis Res. Ther. 11, 1–13.

Li, Y., Lee, P.Y., Kellner, E.S., Paulus, M., Switanek, J., Xu, Y., Zhuang, H., Sobel, E.S., Segal, M.S., Satoh, M., et al. (2010). Monocyte surface expression of Fcγ receptor RI (CD64), a biomarker reflecting type-I interferon levels in systemic lupus erythematosus. Arthritis Res. Ther. 12, 1–12.

Ludvigsson, J.F. (2020). Systematic review of COVID-19 in children shows milder cases and a better prognosis than adults. Acta Paediatr. Int. J. Paediatr. 1088–1095.

Maeno, N., Takei, S., Masuda, K., Akaike, H., Matsuo, K., Kitajima, I., Maruyama, I., and Miyata, K. (1998). Increased Serum Levels of Vascular Endothelial Growth Factor in Kawasaki Disease. Pediatr. Res. 44, 596–599.

Maurer, M., and Von Stebut, E. (2004). Macrophage inflammatory protein-1. Int. J. Biochem. Cell Biol. 36, 1882–1886.

Mirrakhimov, A. (2015). Antisynthetase Syndrome: A Review of Etiopathogenesis, Diagnosis and Management. Curr. Med. Chem. 22, 1963–1975.

Mohan, T., Deng, L., and Wang, B. (2020). International Immunopharmacology CCL28 chemokine?: An anchoring point bridging innate and adaptive immunity.

Van Der Molen, R.G., Hamann, D., Jacobs, J.F.M., Van Der Meer, A., De Jong, J., Kramer, C., Strengers, P.F.W., and Van Der Meer, J.W.M. (2015). Anti-SSA antibodies are present in immunoglobulin preparations. Transfusion (Paris) 55, 832–837.

Nakamura, Y., Yashiro, M., Uehara, R., Sadakane, A., Chihara, I., Aoyama, Y., Kotani, K., and Yanagawa, H. (2010). Epidemiologic features of Kawasaki disease in Japan: Results of the 2007-2008 nationwide survey. J. Epidemiol. 20, 302–307.

Nishimoto, N., Terao, K., Mima, T., and Nakahara, H. (2008). Mechanisms and pathologic significances in increase in serum interleukin-6 (IL-6) and soluble IL-6 receptor after administration of an anti–IL-6 receptor antibody,. Blood 112, 3959–3965.

Onouchi, Y. (2018). The genetics of Kawasaki disease. Int. J. Rheum. Dis. 21, 26–30.

Pietschmann, P., Stohlawetz, P., Brosch, S., Steiner, G., Smolen, J.S., and Peterlik, M. (1998). The effect of alendronate on cytokine production, adhesion molecule expression, and transendothelial migration of human peripheral blood mononuclear cells. Calcif. Tissue Int. 63, 325–330.

van der Poel, C.E., Spaapen, R.M., van de Winkel, J.G.J., and Leusen, J.H.W. (2011). Functional Characteristics of the High Affinity IgG Receptor, FcγRI. J. Immunol. 186, 2699–2704.

Rauf, A., Vijayan, A., and John, S.T. Multisystem inflammatory syndrome with features of Atypical Kawasaki disease during COVID-19 pandemic?: Report of a case from India. 3–7.

Riphagen, S., Gomez, X., Gonzalez-Martinez, C., Wilkinson, N., and Theocharis, P. (2020). Hyperinflammatory shock in children during COVID-19 pandemic. Lancet Lond. Engl. 395, 1607–1608.

La Scola, B., Le Bideau, M., Andreani, J., Hoang, V.T., Grimaldier, C., Colson, P., Gautret, P., and Raoult, D. (2020). Viral RNA load as determined by cell culture as a management tool for discharge of SARS-CoV-2 patients from infectious disease wards. Eur. J. Clin. Microbiol. Infect. Dis. 39, 1059–1061.

Sheikh, N.A., and Jones, L.A. (2008). CD54 is a surrogate marker of antigen presenting cell activation. Cancer Immunol. Immunother. 57, 1381–1390.

Stanley, E.R., and Chitu, V. (2014). CSF-1 receptor signaling in myeloid cells. Cold Spring Harb. Perspect. Biol. 6, 1–21.

Tanaka, M., Krutzik, S.R., Sieling, P.A., Lee, D.J., Rea, T.H., and Modlin, R.L. (2009). Activation of FcγRI on Monocytes Triggers Differentiation into Immature Dendritic Cells That Induce Autoreactive T Cell Responses. J. Immunol. 183, 2349–2355.

Toubiana, J., Poirault, C., Corsia, A., Bajolle, F., Fourgeaud, J., Angoulvant, F., Debray, A., Basmaci, R., Salvador, E., Biscardi, S., et al. (2020). Kawasaki-like multisystem inflammatory syndrome in children during the covid-19 pandemic in Paris, France: prospective observational study. BMJ 369, m2094.

Turnier, J.L., Anderson, M.S., Heizer, H.R., Jone, P.N., Glodé, M.P., and Dominguez, S.R. (2015). Concurrent respiratory viruses and Kawasaki disease. Pediatrics 136, e609–e614.

Uehara, R., and Belay, E.D. (2012). Epidemiology of kawasaki disease in Asia, Europe, and the United States. J. Epidemiol. 22, 79–85.

Vahidy, F.S., Nicolas, J.C., Meeks, J.R., Khan, O., Jones, S.L., Masud, F., Sostman, H.D., Phillips, R.A., Andrieni, J.D., Kash, B.A., et al. (2020). Racial and Ethnic Disparities in SARS-CoV-2 Pandemic: Analysis of a COVID-19 Observational Registry for a Diverse U.S. Metropolitan Population. MedRxiv 2020.04.24.20073148.

Verdoni, L., Mazza, A., Gervasoni, A., Martelli, L., Ruggeri, M., Ciuffreda, M., Bonanomi, E., and Antiga, L.D. (2020). Articles An outbreak of severe Kawasaki-like disease at the Italian epicentre of the SARS-CoV-2 epidemic?: an observational cohort study. The Lancet 6736, 1–8.

Vilgelm, A.E., and Richmond, A. (2019). Chemokins modulate immune surveillance in tumorignesis, metastatsis, and response to immunotherapy. Front. Immunol. 10, 6–8.

Whittaker, E., Bamford, A., Kenny, J., Kaforou, M., Jones, C.E., Shah, P., Ramnarayan, P., Fraisse, A., Miller, O., Davies, P., et al. (2020). Clinical Characteristics of 58 Children With a Pediatric Inflammatory Multisystem Syndrome Temporally Associated With SARS-CoV-2. JAMA 1–11.

WHO (2020). Multisystem inflammatory syndrome in children and adolescents with COVID-19. 1–3.

Williams, I.R. (2006). CCR6 and CCL20: Partners in intestinal immunity and lymphorganogenesis. Ann. N. Y. Acad. Sci. 1072, 52–61.

Wölfel, R., Corman, V.M., Guggemos, W., Seilmaier, M., Zange, S., Müller, M.A., Niemeyer, D., Jones, T.C., Vollmar, P., Rothe, C., et al. (2020). Virological assessment of hospitalized patients with COVID-2019. Nature 581.

Zheng, S., Fan, J., Yu, F., Feng, B., Lou, B., Zou, Q., Xie, G., Lin, S., Wang, R., Yang, X., et al. (2020). Viral load dynamics and disease severity in patients infected with SARS-CoV-2 in Zhejiang province, China, January-March 2020: Retrospective cohort study. The BMJ 369.

Zhou, P., Yang, X. Lou, Wang, X.G., Hu, B., Zhang, L., Zhang, W., Si, H.R., Zhu, Y., Li, B., Huang, C.L., et al. (2020). A pneumonia outbreak associated with a new coronavirus of probable bat origin. Nature 579, 270–273.

Zhu, N., Zhang, D., Wang, W., Li, X., Yang, B., Song, J., Zhao, X., Huang, B., Shi, W., Lu, R., et al. (2020). A novel coronavirus from patients with pneumonia in China, 2019. N. Engl. J. Med. 382, 727–733.

